# SARS-CoV-2 Variant Tracking and Mitigation: Strategies and Results from In-Person Learning at a Midwestern University in the 2020/2021 School Year

**DOI:** 10.1101/2021.08.26.21262704

**Authors:** Carolina Avendano, Aaron Lilienfeld, Liz Rulli, Melissa Stephens, Wendy Alvarez Barrios, Joseph Sarro, Michael E. Pfrender, Marie Lynn Miranda

## Abstract

**Importance:** The COVID-19 pandemic led many higher education institutions to close campuses during the 2020-21 academic year. As campuses prepared for a return to in-person education, many institutions were mandating vaccines for students and considering the same for faculty and staff. This paper demonstrates the effectiveness of such a strategy based on evidence from a mid-sized midwestern university.

**Objective:** To determine whether high vaccination coverage can mitigate the spread of SARS-CoV-2, even in the presence of highly-transmissible variants and congregate living.

**Setting:** This study was conducted at a mid-sized midwestern university during the spring 2021 semester.

**Design:** The university developed a saliva-based surveillance program capable of high-throughput SARS-CoV-2 polymerase chain reaction testing and genomic sequencing with the capacity to deliver results in less than 24 hours. On April 7, 2021, the university announced a vaccine requirement for all students for the fall 2021 semester and announced the same requirement for faculty and staff on May 20, 2021. The university hosted an onsite mass vaccination clinic using the two-dose Pfizer-BioNTech vaccine April 8-15 and April 29 - May 6, 2021. Data from January 6 - May 20, 2021 were analyzed.

**Participants:** This study includes 14,894 individuals from the university population who tested on campus for COVID-19 during the spring 2021 semester.

**Main Outcomes and Measures:** Positive SARS-CoV-2 diagnosis was confirmed by quantitative reverse transcription–polymerase chain reaction (qRT-PCR) of saliva specimens and variant identity was assessed by qRT-PCR and next-generation sequencing (NGS) of viral genomes.

**Results:** Between January and May 2021, the university conducted 196,185 COVID-19 tests and identified 1,603 positives – ∼89% students – with 687 identified via PCR of saliva specimens. The Alpha (B.1.1.7) variant constituted 44% of total positives sequenced. By May 20, 2021, 91% (10,068) of students, 92% (814) of faculty, and 72% (2,081) of staff were vaccinated. The 7-day rolling average of positive cases peaked at 37 cases on February 17 but declined to zero by May 14, 2021. The 7-day moving average of positive cases was inversely associated with the cumulative vaccination rate.

**Conclusions and Relevance:** This study demonstrates the high effectiveness of robust vaccination programs even in the presence of highly-transmissible variants and congregate living.

**KEY POINTS:** *Question:* How is the spread of COVID-19 affected by increasing rates of vaccination?

*Findings:* We leverage 190,000+ COVID-19 surveillance tests including 1,603 positives for COVID-19 at a mid-sized midwestern university from January 6 – May 20, 2021. Genomic sequencing indicates that the Alpha (B.1.1.7) variant was first identified in early February and became the dominant variant by early March and the only variant resulting in positive cases by April. An increase in vaccination was associated with a dramatic decrease in COVID-19 cases in the campus population.

*Meaning:* Mass vaccination efforts can effectively control the spread of SARS-CoV-2 even as highly-transmissible variants are introduced.

## Introduction

After initial nearly universal shutdowns of in-person learning at US colleges and universities in March of 2020, many higher education institutions kept campuses fully or partially closed during the 2020-21 academic year. Those that remained open relied on a variety of public health measures including masking, physical distancing, hand sanitizing stations, ventilation adjustments, plexiglass installation, contact tracing teams, quarantine and isolation facilities, and de-densification.^1^ Some institutions also instituted regular population-level surveillance testing to track and contain the spread of SARS-CoV-2 on campuses.^2-4^ The resources required to launch and maintain such surveillance programs are likely unrealistic for most organizations.^5^ Nevertheless, the data generated from such approaches can provide valuable insight on strategies for mitigating the spread of SARS-CoV-2.

Many US higher education institutions are now mandating vaccination for returning students,^6^ even as vaccine hesitancy remains vexingly high in the country, with many continuing to question the effectiveness and safety of the vaccines. Here we demonstrate how data collected through university surveillance testing and sequencing programs are particularly relevant for assessing the effectiveness of high vaccination coverage in reducing COVID-19 case rates.

## Setting, Design, and Methods

We report on a mid-sized Midwestern university with a total (faculty, staff, and students) population of roughly 16,000, 86% of classes held in-person, and full density in residence halls. The university is located in a mid-sized town of ∼100,000. We focus on the spring 2021 semester.

The university established an in-house CLIA-certified testing facility with the ability to detect SARS-CoV-2 via reverse transcription-polymerase chain reaction testing and genetically analyze samples for the presence and identity of genetic variants. From January 6, 2021 – May 20, 2021, the university conducted 196,185 surveillance tests. For the spring 2021 semester, the university required all undergraduates and professional students to be tested once per week, with every other week voluntary testing for graduate students, faculty and staff.

The surveillance testing program relied on both commercial (61,256 tests; 916 positives) and in-house RT-PCR molecular testing (134,929 tests; 687 positives) for SARS-CoV-2 (see methods in Supplement). Using the saliva specimens collected in-house, genetic variants of SARS-CoV-2 on 446 samples were determined by next generation sequencing (NGS) the viral genomes and comparison to the extensive databases of known variants.^7, 8^

The study population is limited to individuals who were tested for SARS-CoV-2 at least once at the university between January 6, 2021 and May 20, 2021. Descriptive statistics and graphical analysis comparing trends in cases with trends in vaccination were developed using R. The study was reviewed by the university’s institutional review board and was classified as exempt for approval and informed consent.

## Results and Discussion

Between January and May 2021, the university conducted 196,185 COVID-19 tests and identified 1,603 individuals who were positive for SARS-CoV-2, including 1,426 students (90%), 152 staff (10%), and 15 faculty (1%). Samples from the start of the semester showed only the B.1.2 variant. By the end of January, variants designated by the CDC as variants of interest and of concern began to appear on campus.^9^ The Epsilon (B.1.427/B.1.429) and Alpha (B.1.1.7) variants were among the first detected. The Epsilon variant quickly rose in frequency to become the dominant variant in mid-February. Within 5 weeks, the Alpha (B.1.1.7) variant was the majority strain and by 8 weeks after detection accounted for over 90% of cases (See Figure 1). These results elucidate the importance of genomic surveillance in quickly detecting the appearance of VOC among the campus population in order to swiftly implement new mitigating strategies as needed.^10^

**Figure 1.**
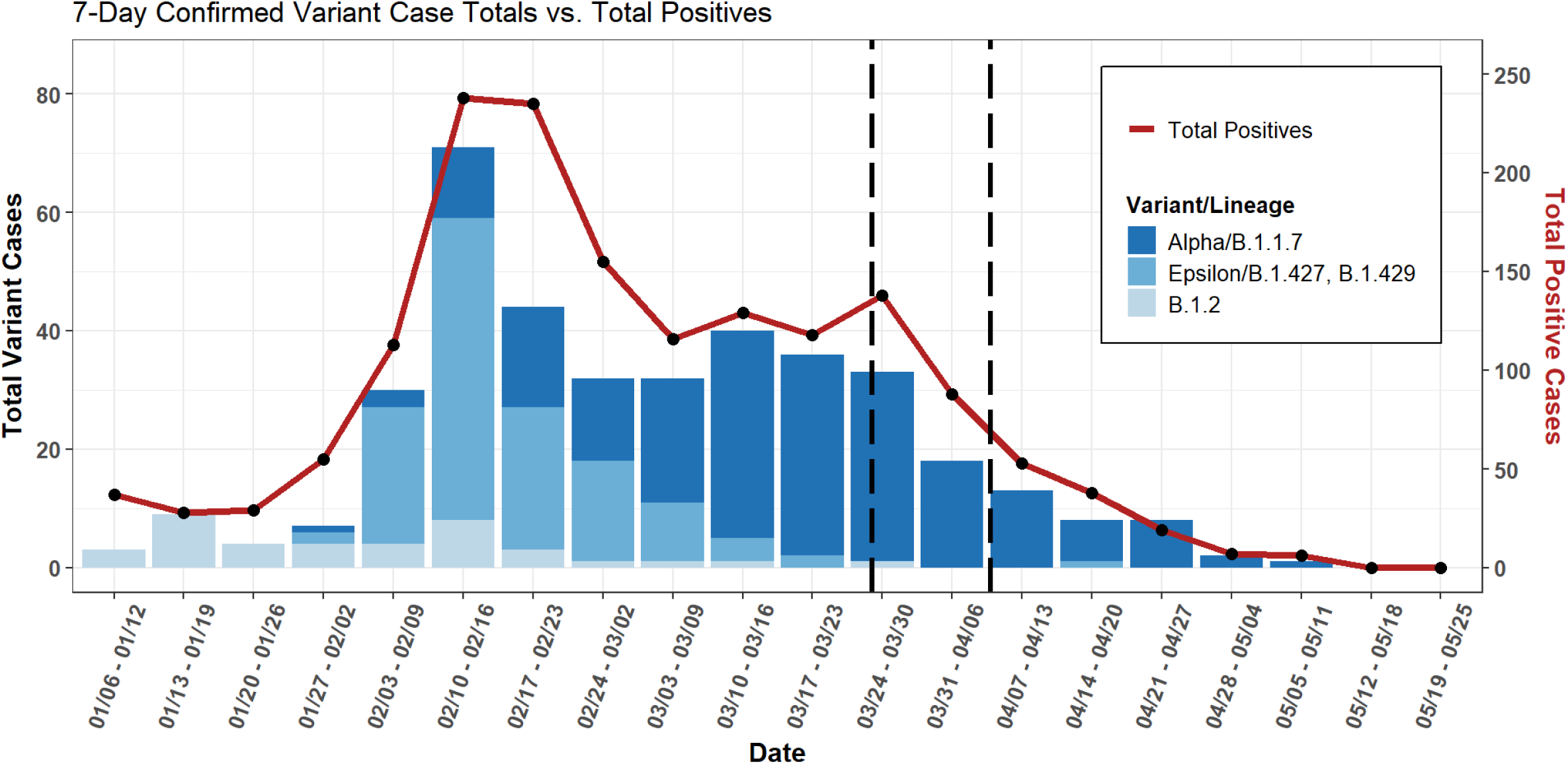
Longitudinal Trend of Positives and Confirmed Variants. 7-Day confirmed Variant Case Totals vs. Total Positives. The first vertical dotted line indicates the first date of the vaccination clinic for the local community, March 27, and the second vertical dotted line indicates the first date of the vaccination clinic for the university population, April 8.

Running parallel to the surveillance testing effort at the university was the increased availability of COVID-19 vaccines. The home state of the university began offering vaccinations on December 14, 2020 first exclusively to health care workers and then with a focus on the older population. By March 31, eligibility for vaccination opened up to all those over the age of 16. By March 31, 2021, 15.4% of individuals 18 and older in the state had received vaccinations.^11^

In an effort to rapidly increase the vaccination rate across the home state, local and state governments partnered with local organizations to host mass vaccination clinics. The university hosted a mass vaccination clinic (Johnson & Johnson) for northern state residents on March 26-27, which provided ready access to vaccines for all county residents, including university faculty and staff. At this clinic, more than 5,000 state residents received vaccines.

Shortly thereafter in April 8, the state provided the university with sufficient Pfizer-BioNTech two-dose vaccines to allow all members of the university community to obtain vaccines, including family members of employees, during an on-campus mass vaccination event. Figure 2 shows that as a result of local and state vaccination efforts, by the start of the university’s mass vaccination campaign on April 8, 50% of the faculty and 30% of the campus population was already fully vaccinated.

**Figure 2.**
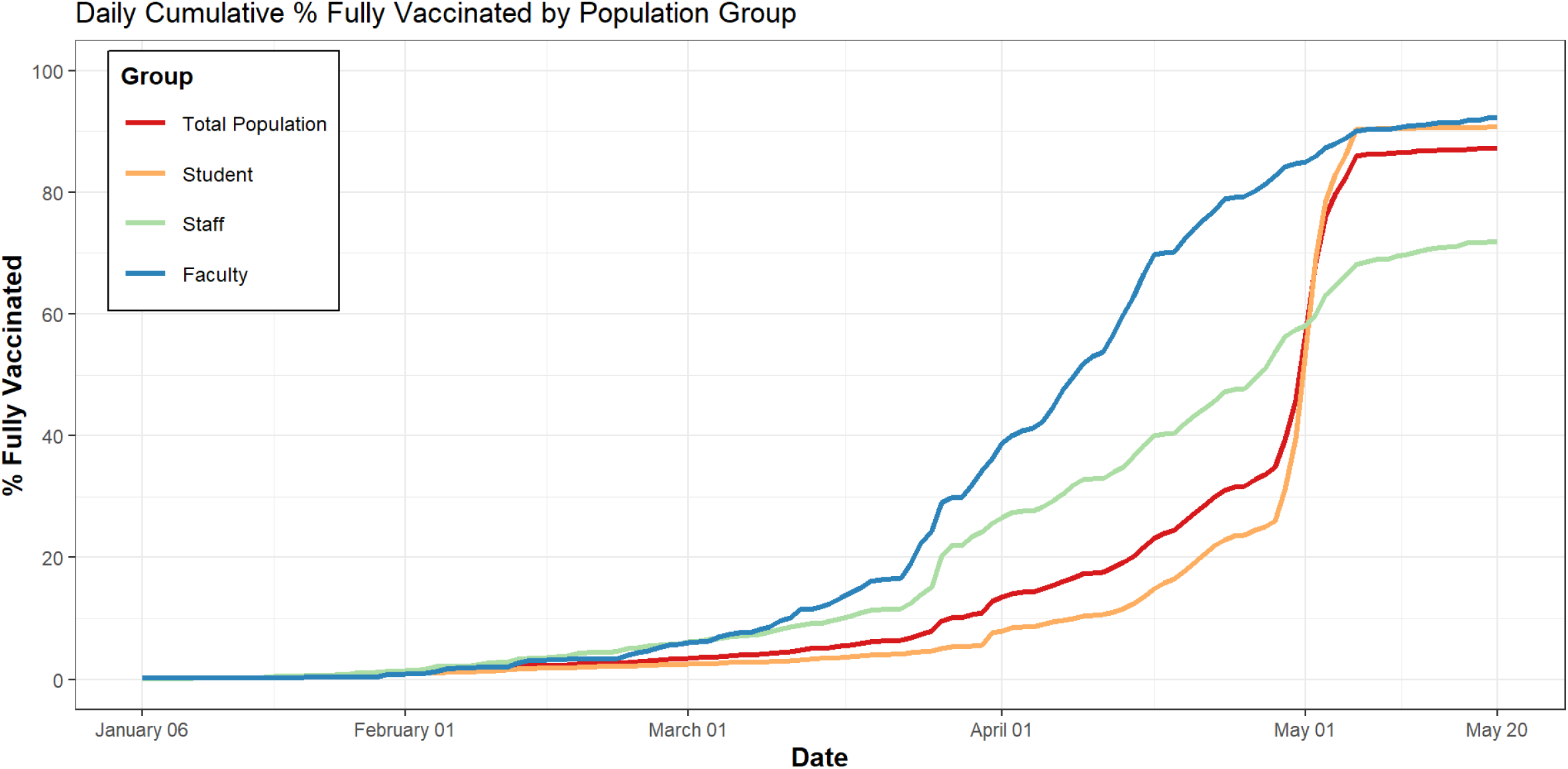
Daily Cumulative % of Fully Vaccinated Individuals by Population Group. The red curve represents the daily cumulative % of fully vaccinated individuals of the total university Spring 2021 in-person population; the yellow curve represents the rate of vaccination for the university Spring 2021 student population; the green curve represents the rate of vaccination for university Spring 2021 staff population; and the blue curve represents the rate of vaccination for the university Spring 2021 faculty population

On April 7, university administration informed the student body that vaccines would be required to enroll during the fall 2021 semester, and offered easing of public health restrictions during graduation weekend if the student body achieved a 90%+ vaccination rate. By May 20, 13,002 individuals from the university community were vaccinated including 10,068 students (91% of the students), 814 faculty (92% of the faculty) and 2,081 staff (72% of the staff). The 91% student vaccination rate compares to 28% of 18-24 year old’s fully vaccinated nationwide^12^ and 32% statewide^13^ by early May 2021. By May 20, 87 % of the total in-person campus population had been fully vaccinated compared to 53% of the 18 and older population in the local county (See Figure 2).^14^

Within two weeks after the first dose of the Pfizer BioNtech vaccine, positive cases fell to their lowest levels of the semester, and continued to decrease for the remainder of the school year achieving 2 weeks of zero cases by the end of the semester. Just as the second dose was administered, the positivity rate via surveillance testing decreased from 1% to 0.1% (see eFigure 1 in Supplement). In contrast, the local county incidence rate remained flat during the same period of time which suggests that the campus population vaccination rate played a role in the rapid decrease of positive cases on campus (see Figure 3).

**Figure 3.**
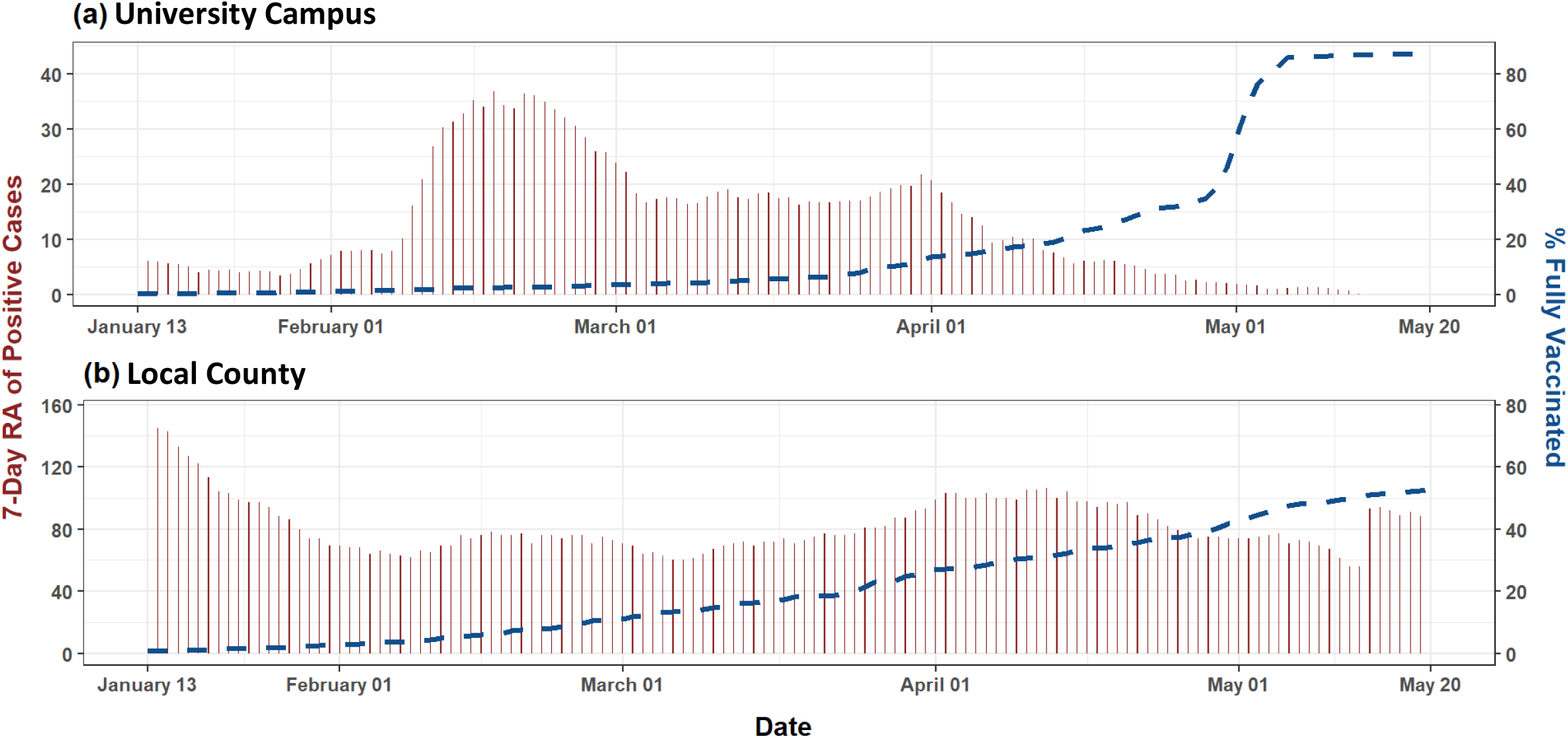
Longitudinal Trend of Positives and Percent Vaccinated in the Population. Comparison of (a) university campus and (b) local county 7-Day rolling average of positive cases versus the percent of fully vaccinated

Figure 3 shows the % fully vaccinated versus the 7-day moving average positivity rate. The 7-day moving average of positive cases was inversely associated with the cumulative vaccination rate, with a statistically significant correlation of −0.57 (95% CI = −0.68, −0.44). A robust vaccination program was clearly associated with a reduction in COVID-19 cases.

## Conclusions

While weekly surveillance testing as practiced by some colleges and universities to contain the spread of COVID-19 is well beyond the scope of many organizations, data from these efforts are useful for other purposes. At a mid-sized Midwestern university, which had previously exhibited persistent and significant numbers of COVID-19 cases despite robust public health protocols, high vaccination coverage interrupted the prevailing transmission patterns. This was true despite predominantly in-person education, full density congregate living, and the presence of the more transmissible B.1.1.7 (alpha) variant. These results support the decision by many colleges and universities to require vaccines for the fall semester and more generally provide evidence of the efficacy of COVID-19 vaccines.

## Data Availability

Datasets are available from the corresponding author upon reasonable request. All datasets have restricted access and are governed by an IRB at the University of Notre Dame.

## Author Contributions

Carolina Avendano and Aaron Lilienfeld had full access to all of the data in the study and take full responsibility for the integrity and the accuracy of the data analysis.

*Concept and design:* Avendano, Lilienfeld, Miranda, Pfrender, Rulli

*Acquisition, analysis, or interpretation of data:* Avendano, Lilienfeld, Pfrender, Rulli, Sarro, Stephen,

*Drafting of the manuscript:* Avendano, Lilienfeld, Miranda, Pfrender, Rulli

*Critical revision of the manuscript for important intellectual content:* Avendano, Miranda, Pfrender

*Statistical analysis:* Lilienfeld, Miranda

*Administrative, technical, or material support:* Avendano, Barrios, Rulli, Stephens

*Supervision:* Miranda, Pfrender, Rulli

## Additional Contributions

The University of Notre Dame COVID Response Unit supported data collection. Claire Osgood provided data management expertise. All acknowledged individuals were paid by ND for their efforts and have provided written consent to being named in this publication.

