## Supplemental Material for "SARS-CoV-2 Variant Tracking and Mitigation: Strategies and Results from In-Person Learning at a Midwestern University in the 2020/2021 School Year"

**eMethods.** Saliva Sample Preparation, RNA Extraction, Illumina Whole Genome Library Preparation and Sequencing, and NGS Data Analysis and Variant Determination.

**eFigure 1.** Positive After Vaccination

### eMethods

#### Saliva Sample Preparation

Fresh saliva samples collected from study participants were heat inactivated at 95°C for 30 min. Following the steps outlined by Ranoa *et al.*[1], 2020, samples were diluted 1:1 (vol/vol) with 2XTris-Borate-EDTA buffer (0.089M Tris, 0.089M Borate, 0.002M EDTA in final 1x buffer solution), and tested for the presence of SARS-CoV-2 via RT-qPCR using the TaqPath COVID-19 Combo kit with TaqPath 1-Step Master Mix (Thermo Fisher Scientific, Cat# A47814, A28523). Following analysis, all heat inactivated positive samples were vortexed lightly, aliquoted into nuclease-free 1.5 mL tubes, and frozen at -80°C for further processing.

#### RNA Extraction

Saliva samples selected for extraction were identified as SARS-CoV-2 positive from the RT-qPCR assay with Ct values ≤32. These Ct values resulted from the combined average of the SARS-CoV-2 gene targets (N-gene, S-gene, ORF1ab) that were collected from two independent tests. Frozen saliva samples were thawed on ice, lightly vortexed, and pulse centrifuged for 12 seconds to a maximum speed of 1000 rpm to pellet suspended debris and solid particles. Viral RNA was then extracted from 140ul of saliva using the QIAamp Viral RNA Mini Kit according to the manufacturer's instructions for spin columns (Qiagen, Inc). RNA was eluted in 20-40 µl of nuclease free molecular grade water and placed at -80°C for storage until ready for library preparation.

#### Illumina Whole Genome Library Preparation and Sequencing

RNA isolated from saliva samples was used as the starting material for library preparation using the Illumina COVIDSeq Test (Illumina Inc.), according to the manufacturer's instructions. Library preparation and sequencing was performed at the Genomics & Bioinformatics Core Facility at Notre Dame. Briefly, cDNA was synthesized from 8.5ul extracted RNA and amplified using two separate primer pools tiling the SARS-Cov-2 genome. Following PCR, the two amplicon pools were combined and a bead-linked transposome fragmented and tagged the amplicons with adapter sequence. Libraries were then indexed and pooled by volume. The pooled libraries were quality assessed using a combination of Qubit dsDNA HS Assay (Thermo Fisher Scientific), Bioanalyzer DNA 1000 Assay (Agilent Technologies), and KAPA Library Quantification Kit for Illumina (Kapa Biosystems), and normalized to 4 nM to prepare for sequencing. Library pools were sequenced on Illumina NextSeq 500 High Output v2.5 (300 cycle) flowcells using paired-end 2x150bp reads to a minimum read depth of 1 Million reads per sample.

#### NGS Data Analysis and Variant Determination

Raw sequences were assessed for quality with FastQC version v0.11.8.[2] Information, including read depth, was extracted from the FastQC output using custom scripts.[3] FASTQ files were processed using the DRAGEN COVID Lineage app (Illumina Inc.) This app built FASTA files, established clade identity through Nextclade[4], and lineages identity through Pangolin.[5] Clade and lineage assessments were verified through independent processing using Nextclade and Pangolin respectively. Results were deposited to GISAID[6] and Genbank.[7] Submission files were generated with custom scripts.[3]

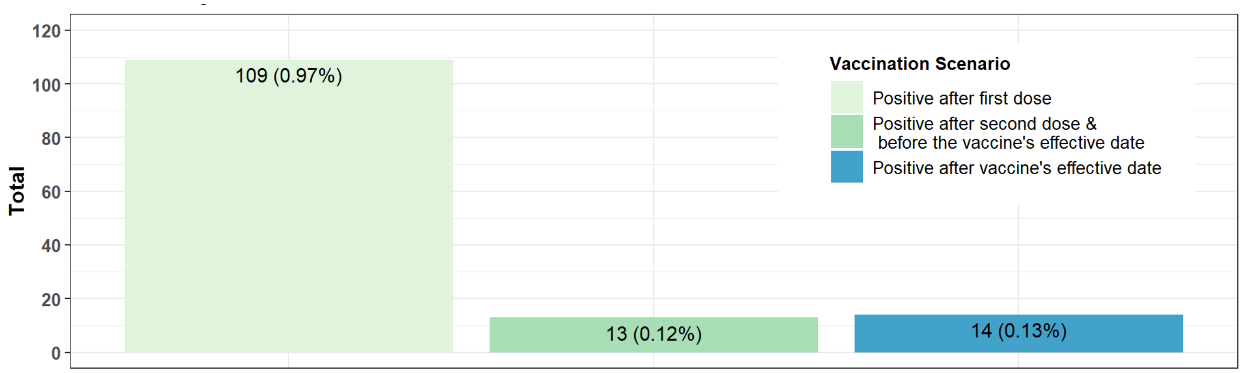


**eFigure 1. Positive After Vaccination.** Percentages represent the proportion of those positive after vaccination over the total Spring 2021 population after first dose, immediately after second dose, and after fully vaccinated (two weeks after second dose).
